# Multimodal neuroimaging approach for cognitive impairment in Alzheimer’s disease

**DOI:** 10.64898/2026.06.04.26354924

**Authors:** Marlon Gonzales, Xiaojian Kang, Maheen M. Adamson, Steven Z. Chao, Byung C. Yoon, the Alzheimer’s Disease Neuroimaging Initiative

## Abstract

**PURPOSE:** Alzheimer’s disease (AD) is associated with cognitive impairment, brain atrophy, and elevated amyloid-beta (Aß) and tau. The study aimed to characterize regional atrophy associated with elevated Aß and tau, as measured by [¹ F]florbetapir (FBP) and [¹ F]flortaucipir (FTP) positron emission tomography (PET), respectively, and determine whether combining PET and atrophy data improves the prediction of cognitive impairment.

**METHODS:** Alzheimer’s Disease Neuroimaging Initiative data (n = 381) were retrospectively analyzed. PET results were correlated with cortical thickness, gray matter (GM) volumes, Mini-Mental State Examination, and Montreal Cognitive Assessment. Linear/logistic regression and area under the curve (AUC) were used to evaluate for significant correlations and compare performances in distinguishing cognitive impairment, respectively.

**RESULTS:** Incremental loss of cortical thickness and GM volume was observed from FBP-/FTP-(n = 205) to single PET-positive (FBP+/FTP-, n = 133; FBP-/FTP+, n = 5) and FBP+/FTP+ (n = 38) groups, particularly in the temporal and parietal lobes. FBP+/FTP+ showed the most severe cortical thickness loss in the entorhinal cortex, temporal lobe GM atrophy, and cognitive impairment. Adding brain atrophy as the third variable resulted in higher odds ratios and improved AUCs for cognitive impairment, with FBP+/FTP+/temporal GM or entorhinal cortical atrophy+ demonstrating the strongest associations with cognitive impairment.

**CONCLUSION:** A multimodal approach combining PET and MRI may help improve the assessment of cognitive impairment in AD.

## INTRODUCTION

Alzheimer’s disease (AD) is a neurodegenerative disease characterized by progressive cognitive decline [1]. It is the leading cause of dementia, with the incidence and prevalence doubling every 5 years after the age of 60 [2]. It is estimated that 13.8 million Americans aged 65 and older may be diagnosed with AD by 2060 [3]. The current diagnostic criteria include abnormal cerebrospinal fluid or positron emission tomography (PET) studies using amyloid-beta (Aβ) tracers [1]. Two popular PET imaging tracers that detect abnormal biomarkers are [¹ F]florbetapir (FBP), which binds to Aβ plaques in the brain [4] and [¹ F]flortaucipir (FTP), which targets tau accumulation associated with AD [5,6].

Elevation in PET tracers has been linked to brain atrophy and cognitive decline [7,8]. Tau is thought to be more strongly associated with regional brain atrophy than Aβ [9,10] and is believed to be a better predictor of cognitive decline [10,11]. Tau has been associated with widespread gray matter (GM) volume loss affecting all four lobes of the brain [12]. In particular, among patients with mild cognitive impairment or AD, elevated FTP in the entorhinal cortex was associated with lower GM density in the mesial temporal lobe [12]. Aβ has been correlated with atrophy of various brain regions in the temporal lobe, posterior cingulate cortex, and precuneus, with a stronger association with cortical thickness reduction than with volumetric loss [13]. However, other studies have found no significant or weak correlations between Aβ-positivity and regional brain atrophy [10,14].

Despite the growing literature, there is limited understanding of how the patterns of cortical thickness and brain volume loss differ in individuals with FBP and/or FTP positivity. In particular, there is a relative paucity of data on individuals who are Aβ-negative but tau-positive (FBP-/FTP+), as they represent only 4.1% of the cognitively unimpaired population [15]. Furthermore, previous studies have typically analyzed either cortical thickness or brain volumes in isolation [7,10,14,16]. There is also no specific brain region and regional atrophy cut-off established to be clinically relevant for predicting cognitive impairment. Lastly, it is not fully understood to what extent individual imaging modality (i.e. FBP, FTP, or MRI) can predict cognitive impairment and whether the combined use of multiple modalities is superior to a single modality.

Using the Alzheimer’s Disease Neuroimaging Initiative 3 (ADNI 3) data, this study aims to determine specific associations between FBP, FTP, and regional brain atrophy as well as correlations between individual neuroimaging modalities and cognitive impairment.

## METERIALS AND METHODS

### Participants

Data used in the preparation of this article were obtained from the ADNI database (adni.loni.usc.edu) [17]. The ADNI was launched in 2003 as a public-private partnership, led by Principal Investigator Michael W. Weiner, MD. The primary goal of ADNI has been to test whether serial magnetic resonance imaging (MRI), positron emission tomography (PET), other biological markers, and clinical and neuropsychological assessment can be combined to measure the progression of mild cognitive impairment (MCI) and early Alzheimer’s disease (AD). For up-to-date information, see www.adni-info.org. Among the ADNI-3 cohort, 381 participants with available FBP, FTP, and MRI data were selected for the study. Demographic variables included age, sex, and years of education. Baseline cognitive function was assessed using Mini-Mental State Examination (MMSE) and the Montreal Cognitive Assessment (MoCA). Cognitive impairment was defined as MMSE ≤ 26 [18] and a MoCA < 26 [19]. Relatively high MMSE and MoCA cut-ff scores were selected to include subjects with the early/mild cognitive impairment.

### PET imaging and classification

PET imaging used FBP and FTP that target and image Aβ and tau proteins, respectively. Baseline Standardized Uptake Value Ratio (SUVR) values of FBP and FTP were collected from ADNI-3. FBP positivity was defined as SUVR > 1.1 [20] FTP positivity was calculated based on past studies [21–23]. For each participant, regional FTP SUVR values were averaged across all available brain regions. The FTP SUVR cut-off was calculated as two standard deviations above the mean FTP SUVR of cognitively normal and FBP-individuals. A cut-off value of 1.265326 was determined, above which was classified as FTP+.

### MRI acquisition and analysis

Magnetic resonance imaging (MRI) studies were acquired on GE, Philips, or Siemens 3T MRI scanners using the standardized three-dimensional magnetization-prepared rapid acquisition with gradient echo (3D MP RAGE) sequence per the ADNI protocol (https://adni.loni.usc.edu/data-samples/adni-data/neuroimaging/mri/mri-scanner-protocols/). The images were processed using FreeSurfer 8.0. GM volume and cortical thickness were obtained for the six anatomical lobes and 34 Desikan Killiany atlas parcels from the FreeSurfer pipeline (Supplemental Table 1). ANOVA analysis was performed to examine the variability among different MRI scanners (Matlab2024b).

### Statistical analysis and data visualization

Unless otherwise specified, statistical analyses and data visualization were performed using RStudio 4.4.3. Linear regression models were used to evaluate associations between continuous FBP and FTP SUVR values and regional cortical thickness and GM volume metrics. Beta-coefficient differences for the PET status were normalized through subtraction from the FBP-/FTP-group. ANOVA models were used to compare regional brain measures across the four FBP/FTP groups, with Bonferroni-corrected post hoc tests to determine group-wise differences. Bonferroni-corrected *p-*values were calculated by multiplying individual *p-*values and the number of comparisons (i.e. corrected *p-*value = original *p*-value * number of comparisons). The number of comparisons was 161 for the analyses presented in Figures 1, 2, and 3, and Supplemental Table 3. The number of comparisons was 40 for the analysis presented in Supplemental Table 4. Corrected *p* < .05 was considered to be statistically significant.

**Figure 1.**
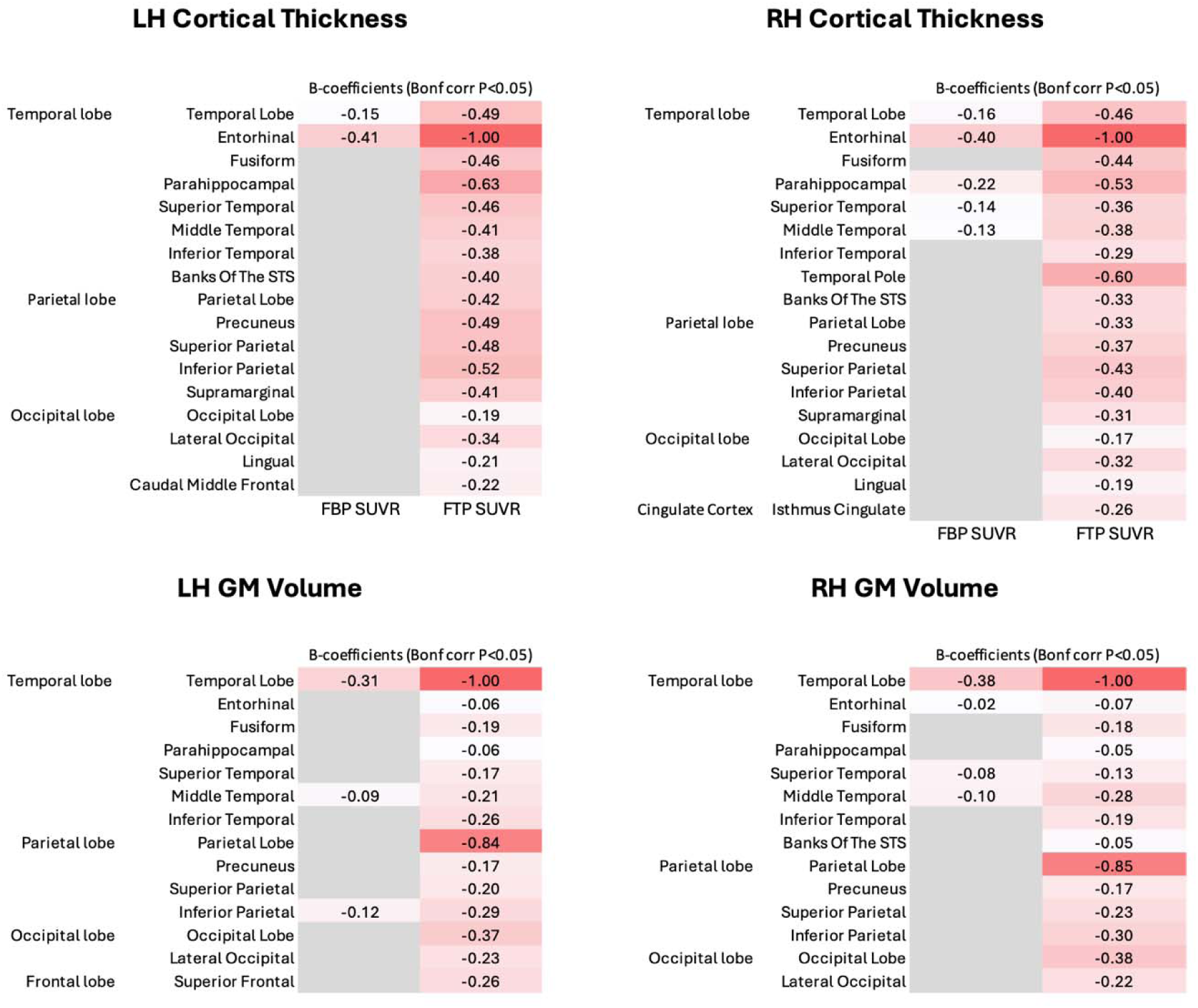
Correlation between FBP/FTP SUVR and regional brain atrophy. Statistically significant (Bonferroni-corrected *p* < .05), normalized beta-coefficients (β) of correlations between various brain regions and FBP/FTP SUVR are listed. For individual brain region, cortical thickness and gray matter volumes were analyzed. Both the left and right hemispheres (LH and RH, respectively) were examined. The beta-coefficients were scaled from −1 to 1. +1 = the most positive beta-coefficient in the regions analyzed (largest magnitude and positive), −1 = the most negative beta-coefficient (largest magnitude and negative), 0 = no effect. FBP, Florbetapir; FTP, Flortaucipir; SUVR, standardized uptake value ratio; STS, superior temporal sulcus.

For relationships between neuroimaging and cognitive measures, binary logistic regression was performed using the MMSE and MoCA cognitive impairment cut-offs as outcomes. Two separate models were created using FBP-/FTP-as a reference to assess cognitive impairment risk across different PET groups, while also including demographic covariates to evaluate their contribution.

Additional logistic regression models incorporated structural atrophy in assessing cognitive impairment risk, with FBP-/FTP-/Atrophy-as the reference group. Atrophy positivity was defined as regional cortical thickness or GM volume one standard deviation below the mean derived from participants from the same reference group the FTP SUVR threshold was calculated from (i.e. cognitively normal and FBP-). Variance inflation factors (VIFs) were calculated to evaluate for multicollinearity of independent variables/predictors (i.e. FBP, FTP, and brain atrophy) and were noted to be relatively low with VIFs ranging from 1.09 to 1.95 (Supplemental Table 2). All logistic regression models were adjusted for TIV, age, sex, and education. The performance of the logistic regression models in distinguishing cognitive impairment was evaluated using the area under the curve (AUC) analysis.

Data were visualized using base plotting functions and the ggplot2 and gt packages. Regional differences across FBP/FTP status groups were visualized using side-by-side bar plots of mean cortical thickness and GM volume regions. 2D brain atrophy maps were generated using the ggseg package and the Desikan-Killiany (DK) atlas.

## RESULTS

### Cohort characteristics

The cohort (n = 381; 200 females, 181 males) was divided into four groups: FBP+/FTP+ (n = 38), FBP+/FTP-(n = 133), FBP-/FTP+ (n = 5), and FBP-/FTP-(n = 205) (Table 1). The FBP-/FTP+ group had the greatest mean age of 77.2 +/- 6.6 (standard deviation) years old, while FBP+/FTP+ had the lowest mean age of 73.9 +/- 8.2 years old. With the exception of FBP-/FTP+, there were more females than males in each group. The most skewed female predominance was observed with FBP+/FTP+, which included 23 females (60.5%) and 15 males. FBP+/FTP+ had the shortest years of education (15.2 +/- 2.5 years) and had the lowest scores on the MMSE and MoCA cognitive assessments. There was no significant difference in TIV between groups (*p* = .529). Additionally, no significant differences were found between the GE, Philips, and Siemens 3T scanners for the cortical thickness (*p* = .08) and TIV volume (*p* = .5) measurements.

**Table 1.**
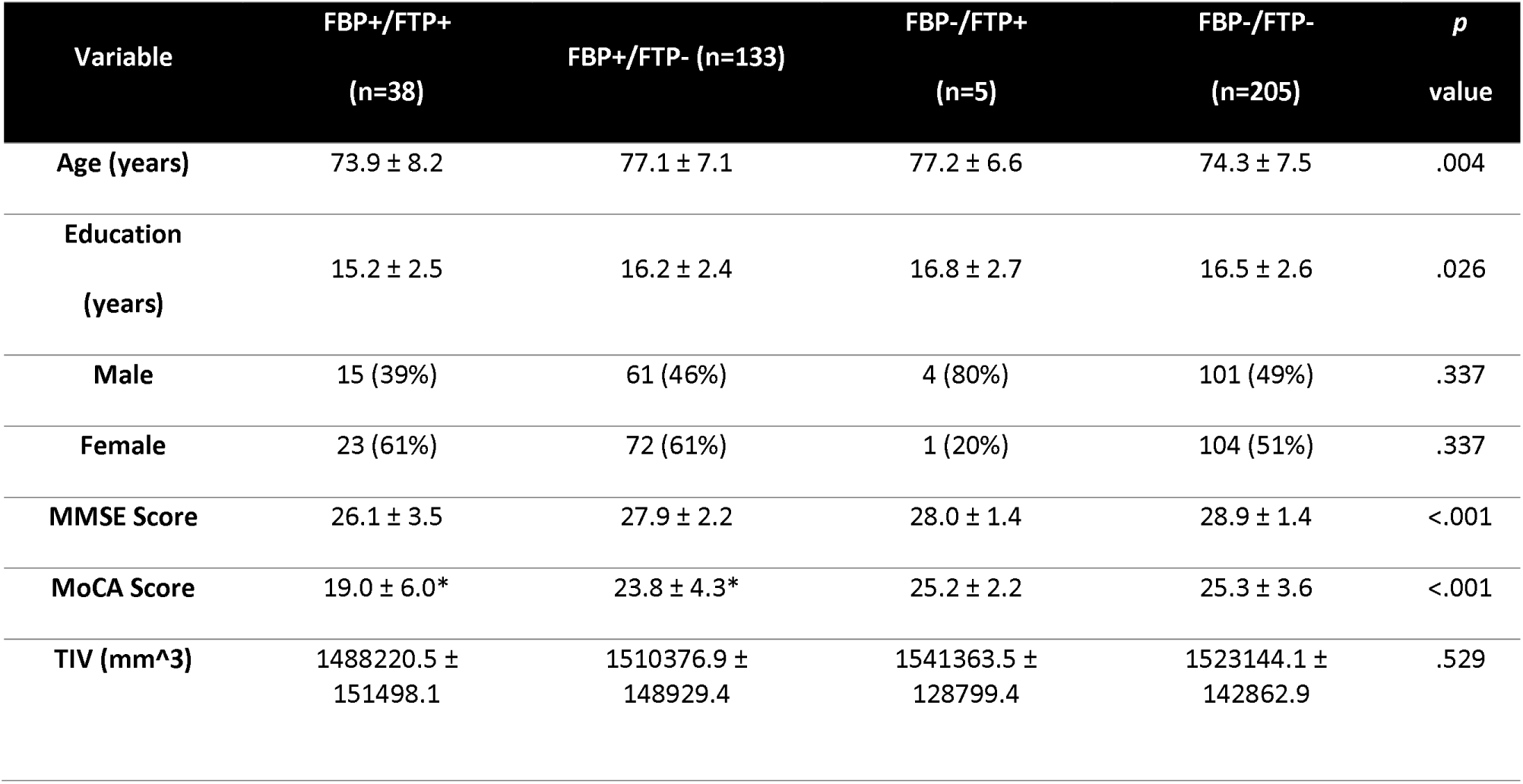
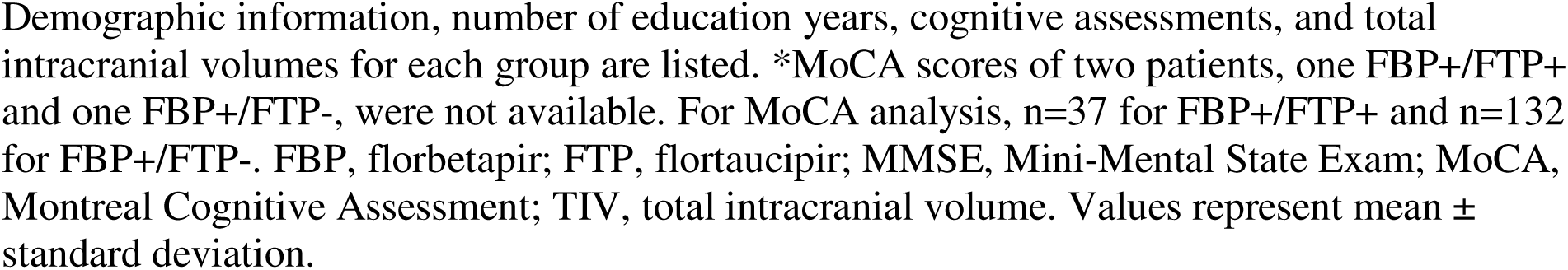
Cohort characteristics.

| Variable | FBP+/FTP+<br>(n=38) | FBP+/FTP- (n=133) | FBP-/FTP+<br>(n=5) | FBP-/FTP-<br>(n=205) | *p*<br>value |
| --- | --- | --- | --- | --- | --- |
| Age (years) | 73.9 ± 8.2 | 77.1 ± 7.1 | 77.2 ± 6.6 | 74.3 ± 7.5 | .004 |
| Education<br>(years) | 15.2 ± 2.5 | 16.2 ± 2.4 | 16.8 ± 2.7 | 16.5 ± 2.6 | .026 |
| Male | 15 (39%) | 61 (46%) | 4 (80%) | 101 (49%) | .337 |
| Female | 23 (61%) | 72 (61%) | 1 (20%) | 104 (51%) | .337 |
| MMSE Score | 26.1 ± 3.5 | 27.9 ± 2.2 | 28.0 ± 1.4 | 28.9 ± 1.4 | <.001 |
| MoCA Score | 19.0 ± 6.0* | 23.8 ± 4.3* | 25.2 ± 2.2 | 25.3 ± 3.6 | <.001 |
| TIV (mm<sup>3</sup>) | 1488220.5 ±<br>151498.1 | 1510376.9 ±<br>148929.4 | 1541363.5 ±<br>128799.4 | 1523144.1 ±<br>142862.9 | .529 |

Demographic information, number of education years, cognitive assessments, and total intracranial volumes for each group are listed. \*MoCA scores of two patients, one FBP+/FTP+ and one FBP+/FTP-, were not available. For MoCA analysis, n=37 for FBP+/FTP+ and n=132 for FBP+/FTP-. FBP, florbetapir; FTP, flortaucipir; MMSE, Mini-Mental State Exam; MoCA, Montreal Cognitive Assessment; TIV, total intracranial volume. Values represent mean ± standard deviation.

### Correlations between PET SUVR values and regional brain atrophy

Direct correlations were made between the regional brain atrophy, as assessed by the cortical thickness and GM volume measurements, and the levels of Aβ and tau, as assessed by FBP and FTP SUVR, respectively. Negative correlations were consistently observed between brain atrophy and the levels of FBP or FTP SUVR, indicating that the increased FBP or FTP levels is associated with decreased brain volumes (or greater atrophy). Between FBP and FTP, FTP SUVR showed substantially more brain regions with statistically significant (Bonferroni-corrected *p* < .05), negative correlations with cortical thickness and GM volume changes than FBP SUVR (Figure 1).

For cortical thickness and FTP SUVR, significant negative correlations were observed in various regions in the temporal lobe, parietal lobe and occipital lobe on both sides as well as the right isthmus of the cingulate (Figure 1). The greatest degree of negative correlations was seen in the temporal lobe (beta-coefficient of −0.49 on the left side and −0.46 on the right; Figure 1), followed by the parietal lobe (−0.42 on the left and −0.33 on the right). More granular regional analysis revealed the greatest negative correlations between the entorhinal cortical thickness and FTP SUVR (−1.00 on both sides). A similar pattern was observed for the GM volume, where the temporal lobe showed the greatest negative correlations (−1.00 on both sides), followed by the parietal lobes (−0.84 on the left and −0.85 on the right).

The correlations between the cortical thickness and FBP SUVR were statistically significant only in the temporal lobes on both sides. In particular, the entorhinal cortex demonstrated the greatest negative correlations (−0.41 on the left and −0.40 on the right). For GM volumes, the temporal lobe GM volumes also showed the greatest negative correlations with FBP SUVR. The left inferior parietal GM volume was the only non-temporal lobe region that showed a significant correlation with FBP SUVR. The correlation between the entorhinal cortex GM volume and FBP SUVR was not significant on the left side and showed significant but minimal, negative beta-coefficient of −0.02 on the right side.

In summary, greater negative correlations were generally observed between cortical thickness/GM volumes and FTP SUVR, compared to FBP SUVR. The temporal lobe cortical thickness and GM volumes showed the highest degree of negative correlations to both FTP and FBP SUVR. The cortical thickness of the entorhinal cortex, but not the GM volume, showed significant negative correlations to FBP SUVR.

### PET positivity and regional brain atrophy

We next examined whether FBP and/or FTP positivity is associated with cortical thickness or GM volume changes within specific brain regions, and whether single or double PET positivity has different degrees of correlation with the regional brain changes.

Significant differences in cortical thickness and GM volume were observed in various brain regions across FBP/FTP groups (*p* < .05; Figure 2, Figure 3, Supplemental Table 3). Most brain regions demonstrated decreased cortical thickness and/or GM volume for FBP+/FTP+, FBP-/FTP+, and FBP+/FTP-, compared to FBP-/FTP-. For cortical thickness, FBP+/FTP+ demonstrated the greatest degree of cortical thickness loss in general. This was followed by FBP-/FTP+, which in turn demonstrated more cortical thickness loss than FBP+/FTP-. For instance, the relative, overall mean cortical thickness compared to FBP-/FTP-was −0.090 mm for FBP+/FTP+, −0.072 mm for FBP-/FTP+, and −0.008 mm for FBP+/FTP-(*p* < .001, Supplemental Table 3). FBP-/FTP+ showed the greatest cortical thickness in select brain regions including the bilateral inferior temporal gyri, bilateral superior temporal gyri, left supramarginal gyrus, right entorhinal cortex, and right temporal lobe (Figure 2 and Figure 3). The entorhinal cortex showed the greatest cortical thickness loss across the FBP/FTP groups (Figure 2). Nearly all of the brain regions with significant differences showed a greater loss of cortical thickness in FBP+/FTP+ and/or FBP-/FTP+, compared to FBP+/FTP-.

**Figure 2.**
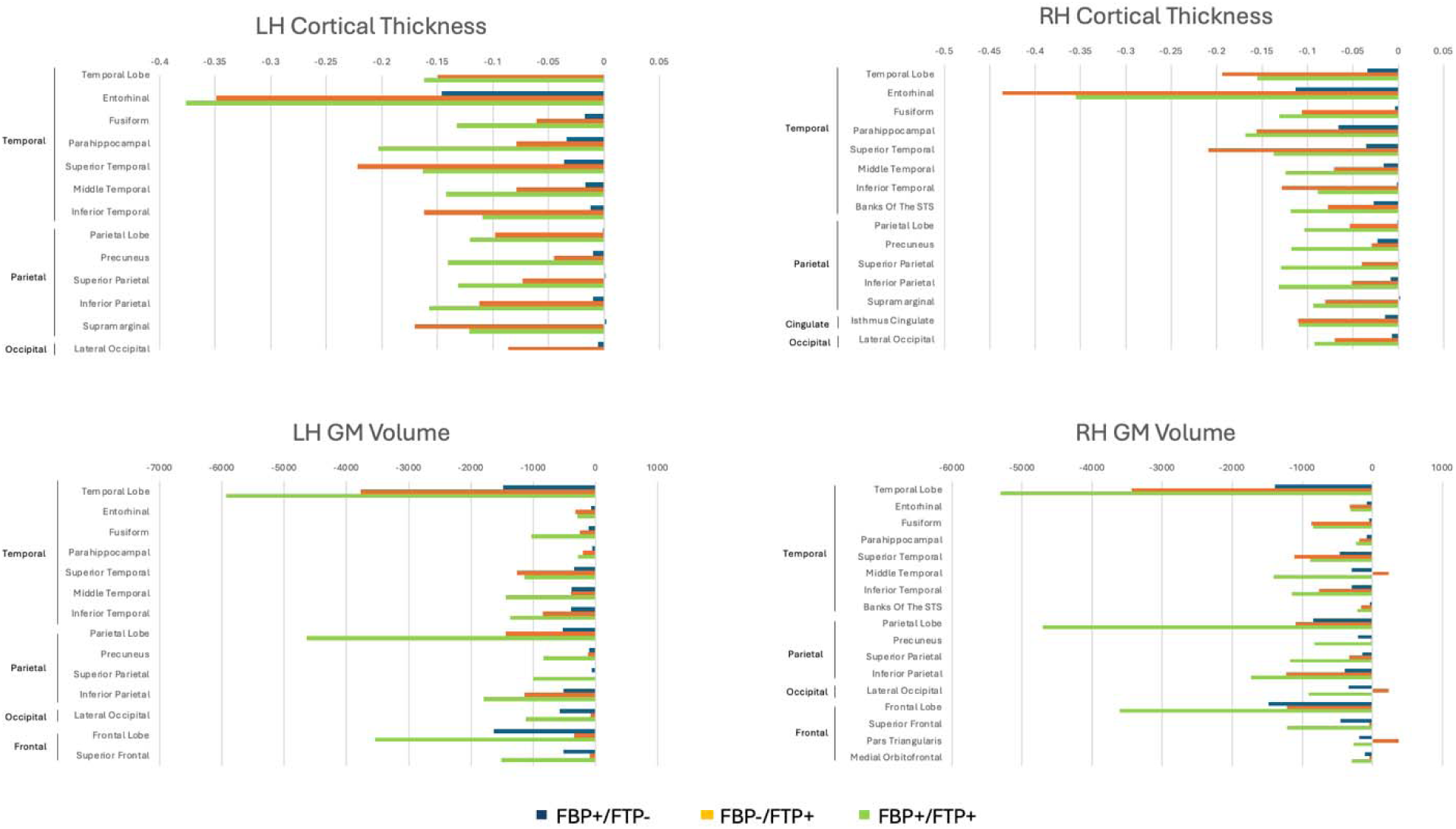
Mean differences in cortical thickness and GM volume compared to FBP-/FTP-group. Bars represent mean regional differences in cortical thickness (mm) and GM volume (×1,000 mm³), normalized to the FBP/FTP group by subtraction. Only the statistically significant (Bonferroni-corrected *p <* .05) differences are presented. Regions without statistically significant group differences (Bonferroni-*corrected p* ≥ .05) in a given hemisphere or measure are omitted from the respective plots (i.e., no bar lines). FBP, Florbetapir; FTP, Flortaucipir; STS, superior temporal sulcus.

**Figure 3.**
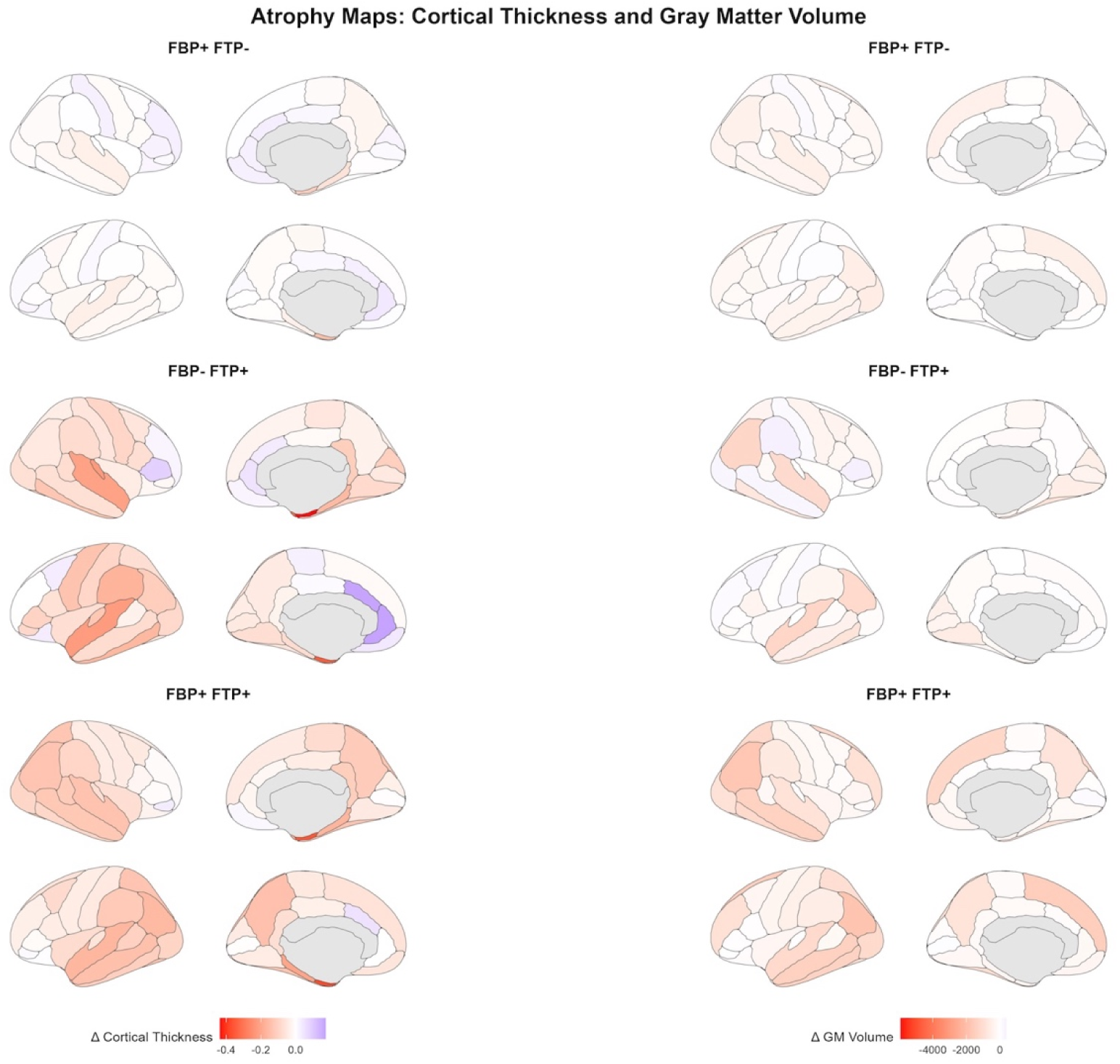
Brain regions with cortical thickness and GM volume changes, compared to the FBP-/FTP-group. Brain atrophy maps were generated based on the Desikan-Killiany (DK) atlas. The maps depict the distribution of statistically significant (Bonferroni-corrected *p* < .05) regional cortical thickness (left) or gray matter volume loss (right) across PET biomarker groups compared to the baseline FBP-/FTP-group. FBP, Florbetapir; FTP, Flortaucipir.

A similar trend was observed with GM volume, where FBP+/FTP+ demonstrated the greatest GM volume loss, followed by FBP-/FTP+ and by FBP+/FTP-. The greatest GM volume changes were seen in the left and right temporal lobes (Figure 2). For instance, FBP+/FTP+ had the GM volume change of −5927 mm^3^ in the left temporal lobe compared to FBP-/FTP-, while FBP-/FTP+ and FBP+/FTP-showed the GM volume changes of −3771 mm^3^ and −1479 mm^3^, respectively (*p* < .001; Figure 2). The second most prominent GM volume loss in FBP+/FTP+ was seen in the parietal lobes in both hemispheres, followed by both frontal lobes (Figure 2). Compared to cortical thickness, GM volumes showed fewer brain regions where FBP-/FTP+ showed more atrophy than FBP+/FTP+ (Figure 2 and Figure 3).

### Associations with cognitive impairment

Associations between various cohort characteristics, including demographics, PET results, and cognitive impairments, were assessed (Table 2). Double positivity in PET assessments (i.e. FBP+/FTP+) exhibited the highest odds of cognitive impairment, with odds ratios (OR) of 10.3 for MMSE ≤ 26 (*p <* .001) and 5.7 for MoCA < 26 (*p* < .001). In comparison, single PET positivity (i.e. FBP+/FTP-) demonstrated a lower OR of 4.7 for MMSE (*p <* .001) and 1.78 for MoCA (*p* = .017). The FBP-/FTP+ group was excluded from this analysis due to the small sample size and the lack of individuals in the group who had a MoCA < 26 or MMSE ≤ 26.

**Table 2.** Associations with cognitive impairment.

| Group (vs FBP- FTP-) | OR | Lower 95% CI | Upper 95% CI | *p* value |
| --- | --- | --- | --- | --- |
| **MMSE $\leq 26$** |  |  |  |  |
| Age (per year) | 0.998 | 0.957 | 1.040 | .917 |
| Male vs Female | 2.800 | 1.210 | 6.680 | .018 |
| Years of Education | 0.878 | 0.771 | 0.997 | .046 |
| TIV | 1.000 | 1.000 | 1.000 | .459 |
| FBP+ FTP- | 4.700 | 2.300 | 10.100 | <.001 |
| **FBP+ FTP+** | **10.300** | **4.200** | **25.900** | **<.001** |
| **MoCA $< 26$** |  |  |  |  |
| Age (per year) | 1.050 | 1.020 | 1.080 | .002 |
| Male vs Female | 2.640 | 1.420 | 4.990 | .002 |
| Years of Education | 0.830 | 0.753 | 0.912 | <.001 |
| TIV | 1.000 | 1.000 | 1.000 | .258 |
| FBP+ FTP- | 1.780 | 1.110 | 2.880 | .017 |
| **FBP+ FTP+** | **5.700** | **2.340** | **16.200** | **<.001** |

Logistic regression models were used to assess the associations between cognitive impairment (i.e. MMSE $\leq 26$ or MoCA $< 26$ ) and various factors including demographics, years of education, total intracranial volume, and FBP/FTP status. FBP, florbetapir; FTP, flortaucipir; OR, odds ratio; CI, confidence interval; MMSE, Mini-Mental State Exam; MoCA, Montreal Cognitive Assessment; TIV, total intracranial volume.

The value of incorporating the regional brain atrophy data as an independent variable was also explored (Table 3 and Supplemental Table 4). For this, the mean cortical thickness of both hemispheres, cortical thickness of the entorhinal cortex, as well as the GM volumes of bilateral temporal and bilateral parietal lobes were classified as positive for atrophy if the respective values were below one standard deviation from the mean values derived from participants who were cognitively normal and PET negative. Mean cortical thickness of both hemispheres was selected given that it represents more global changes in the brain structure. The cortical thickness of the entorhinal cortex was examined given that it was most strongly associated with PET positivity. Similarly, GM volumes of the bilateral temporal and parietal lobes were selected because they showed the most consistent atrophy in correlation with PET positivity. One standard deviation was chosen to balance between having sufficiently different individuals from the normal cohort and having a sufficient number of individuals for analysis. When two standard deviations were defined as the cut-off for substantial atrophy, for instance, there were too few individuals in the atrophy-positive groups for statistical analysis.

**Table 3.** Incremental performance gain of multimodal approach for distinguishing cognitive impairment.

| Outcome | Atrophy Region | Model | AUC | Delta AUC (AUC - Tau AUC) |
| --- | --- | --- | --- | --- |
| MMSE ≤ 26 | Mean Cortical Thickness of Both Hemispheres | Tau Only | 0.712 | 0.000 |
|  |  | Tau + Amyloid | 0.741 | 0.029 |
|  |  | Tau + Atrophy | 0.740 | 0.028 |
|  |  | Tau + Amyloid + Atrophy | 0.754 | 0.042 |
|  | Entorhinal Cortex Thickness | Tau Only | 0.712 | 0.000 |
|  |  | Tau + Amyloid | 0.741 | 0.029 |
|  |  | Tau + Atrophy | 0.74 | 0.028 |
|  |  | Tau + Amyloid + Atrophy | 0.758 | 0.046 |
|  | Temporal GM Volume | Tau Only | 0.712 | 0.000 |
|  |  | Tau + Amyloid | 0.741 | 0.029 |
|  |  | Tau + Atrophy | 0.763 | 0.051 |
|  |  | Tau + Amyloid + Atrophy | 0.777 | 0.065 |
|  | Parietal GM Volume | Tau Only | 0.712 | 0.000 |
|  |  | Tau + Amyloid | 0.741 | 0.029 |
|  |  | Tau + Atrophy | 0.751 | 0.039 |
|  |  | Tau + Amyloid + Atrophy | 0.767 | 0.055 |
| MoCA < 26 | Mean Cortical Thickness of Both Hemispheres | Tau Only | 0.715 | 0.000 |
|  |  | Tau + Amyloid | 0.733 | 0.018 |
|  |  | Tau + Atrophy | 0.714 | -0.001 |
|  |  | Tau + Amyloid + Atrophy | 0.733 | 0.018 |
|  | Entorhinal Cortex Thickness | Tau Only | 0.715 | 0.000 |
|  |  | Tau + Amyloid | 0.733 | 0.018 |
|  |  | Tau + Atrophy | 0.745 | 0.030 |
|  |  | Tau + Amyloid + Atrophy | 0.755 | 0.040 |
|  | Temporal GM Volume | Tau Only | 0.715 | 0.000 |
|  |  | Tau + Amyloid | 0.733 | 0.018 |
|  |  | Tau + Atrophy | 0.741 | 0.026 |
|  |  | Tau + Amyloid + Atrophy | 0.750 | 0.035 |
|  | Parietal GM Volume | Tau Only | 0.715 | 0.000 |
|  |  | Tau + Amyloid | 0.733 | 0.018 |
|  |  | Tau + Atrophy | 0.713 | -0.002 |
|  |  | Tau + Amyloid + Atrophy | 0.734 | 0.019 |

The assessment of areas under the curve (AUCs) for single, double, and triple-positivity in PET/atrophy for cognitive impairment reveals improved AUCs in models incorporating all three modalities. Delta AUCs were calculated by subtracting the single modality AUC values (i.e. flortaucipir (FTP)-only values) from multimodal AUCs. Tau, tau-protein levels as assessed by flortaucipir; amyloid, amyloid- $\beta$ levels as assessed by florbetapir; MMSE, Mini-Mental State Exam; MoCA, Montreal Cognitive Assessment.

Consistently across brain regions, triple positivity (i.e. FBP+/FTP+/Atrophy+) had the highest OR for cognitive impairment (Supplemental Table 4). In particular, the presence of substantial atrophy in the entorhinal cortical thickness and temporal GM volume showed the highest OR for both MMSE ≤ 26 and MoCA < 26, with statistical significance. Conversely, FBP+/FTP+ or FBP+/FTP-individuals without substantial regional brain volume loss showed lower OR for cognitive impairment than those with substantial regional brain volume loss. Among different brain regions, temporal GM volume loss in the FBP+/FTP+ group (FBP+/FTP+/temporal GM atrophy+) exhibited the highest OR of 47.69 (*p* < .001) for MMSE ≤ 26. For MoCA < 26, the FBP+/FTP+ group with the entorhinal cortical thickness loss (FBP+/FTP+/entorhinal cortex atrophy+) exhibited the highest OR of 15.07 (*p* < .05).

The performance gain of incorporating FBP, FTP, and atrophy as independent variables for distinguishing cognitive impairment was further explored using the AUC analysis (Table 3). Compared to the single modality assessment (FTP positivity), the addition of FBP and atrophy generally showed a higher, improved AUC. The only exceptions were the combinations of FTP and mean cortical thickness results and FTP and parietal GM volume that showed decreased AUC. However, the difference was minimal with the changes in AUC of −0.001 and −0.002, respectively. For MMSE ≤ 26, the addition of temporal GM volume loss to the PET results was again shown to make the most substantial improvement in AUC by 0.065 (0.777 vs 0.712). For MoCA < 26 the addition of entorhinal cortical loss showed the greatest improvement in AUC by 0.040 (0.755 vs 0.715).

Another general pattern that was observed is that MMSE results showed stronger associations between PET and atrophy positivity. As an example, for mean cortical thickness, OR was significantly elevated in four of the five groups examined for MMSE ≤ 26 (Supplemental Table 4). In comparison, for MoCA <26, none of the groups showed statistically significant increase in OR. Furthermore, the OR was consistently higher for MMSE (4.04 – 28.47) than MoCA (1.19 – 20.05). A similar pattern was observed for the entorhinal cortical thickness, temporal GM volume, and parietal GM volume.

## DISCUSSION

Our results demonstrate that elevated Aß and tau proteins are independently associated with specific brain regional atrophy and that the elevation in tau has a greater association with brain regional atrophy. Most consistent associations between the cortical thickness and GM volume loss and FBP and/or FTP positivity were observed in the temporal lobe. This was true for both more global measurements of the temporal lobe as a whole, as well as more granular regional measurements within the temporal lobes. For GM volumes, the greatest reduction was seen in the temporal lobe as a whole (Figure 2). For cortical thickness, the entorhinal cortex demonstrated the greatest reduction in PET-positive groups, compared to FBP-/FTP-(Figure 2). These results support the previous findings that the entorhinal cortex is a key region involved in the pathogenesis of AD and is closely associated with the abnormal accumulation of Aß and tau protein [24–26]. For both entorhinal cortical thickness and temporal lobe GM volumes, a progressive increase in respective parenchymal losses was observed, with the single PET-positive groups demonstrating greater atrophy than the PET-negative group but less atrophy than the double PET-positive group. A similar pattern was observed for the other regions in the temporal, parietal, occipital, and frontal lobes where the FBP+/FTP+ generally showed the greatest cortical thickness and GM volume loss among the PET groups (Figure 2 and Figure 3). These findings that the elevations in both Aß and tau on PET are associated with a greater loss of cortical thickness, compared to those with elevated Aß only or negative PET, in cognitively healthy older adults [27].

Between FBP and FTP, our results suggest that FTP is generally associated with a greater loss of cortical thickness and GM volume than FBP. For instance, the entorhinal cortical thickness, temporal GM volume, and parietal GM volume were substantially more reduced in the FBP-/FTP+ group than the FBP+/FTP-group (Figure 2). These results corroborate the previous findings that tau accumulation is associated with increased brain atrophy [9,12,28], while Aβ shows weaker associations with brain volume and thickness [14]. In addition, a prior study that included tau-positive only data found similar findings of greater cortical thickness loss in temporal regions such as the entorhinal cortex and inferior temporal gyrus in FBP-/FTP+ individuals than in FBP+/FTP-individuals [29]. However, it should be noted that the FBP-/FTP+ group in our study is too low (n = 5) to draw conclusions, and more studies are needed to determine cause-and-effect relationships. Furthermore, some of the brain regions such as the bilateral frontal GM and superior frontal GM volumes were more reduced in FBP+/FTP-than FBP-/FTP+, which may also be attributed to the small number of the FBP-/FTP+ group, but also may be due to the regional differences in the sensitivity to the abnormal Aß or tau deposition.

Not every brain region demonstrated congruent results between cortical thickness and GM volume assessments. Cortical thickness demonstrated more mixed results when it came to the correlation with PET positivity. For instance, significant reductions in bilateral frontal GM volumes were seen in PET-positive groups compared to the PET-negative group, but no frontal lobe regions showed significant differences in cortical thickness. In comparison, the cortical thickness of the entorhinal cortex was most substantially affected in correlation with the FBP or FTP positivity, but the correlation was less pronounced for the GM volume of the entorhinal cortex (Figures 1 and 2). The frontal lobe GM volumes were significantly reduced in PET-positive groups, but no significant differences were observed for the cortical thickness of the frontal lobes. These differences may be attributed to the differences in the scale of the cortical thickness (in the order of 2-3 mm) compared to GM volumes (in the order of 1000-70000 mm^3^); it is plausible that the cortical thickness measurements may also be more susceptible to the differences in MRI acquisition and image analyses.

For the assessment of cognitive impairment, previous studies have shown that double positivity in Aβ and tau biomarkers is associated with more rapid cognitive decline and disease progression [10,30]. Here, we introduced the third independent variable, brain atrophy, by creating a binary classification of the presence or absence of substantial, regional brain atrophy to assess the risk of cognitive impairment. Among the various brain regions, the entorhinal cortical thickness and temporal lobe GM volume again appeared to be most strongly associated with higher OR and AUC for cognitive impairment as measured by MMSE or MoCA. These results corroborate the prior findings that the combination of PET data and MRI data, the entorhinal cortical thickness in particular, was additively associated with memory impairment [24]. Our results suggest that more global measurements of mean cortical thickness as well as lobar gray matter volumes could also be used potentially to augment assessment and risk stratification of cognitive impairment.

Interestingly, MMSE results were more strongly correlated with PET or brain atrophy positivity, compared to MoCA. In comparison, previous studies comparing the accuracy of MMSE and MoCA for the assessment of mild cognitive impairment and AD showed the superiority of MoCA over MMSE. A systematic review by Pinto *et al.*, for instance, revealed that more than 80% of the 34 articles they examined showed MoCA to be superior to MMSE in the ability to identify mild cognitive impairment [31]. For various brain regions, a greater number and degree of elevated OR were observed when the regional atrophy data were incorporated for MMSE ≤ 26, compared to MoCA <26 (Table 3). It is possible that MoCA is more sensitive for the detection of cognitive impairment and may identify cognitive impairment at an earlier disease stage. This may be supported by the observation that MoCA scores were consistently lower compared to MMSE across different groups, including the PET negative group that had the mean MoCA score of 25.3 +/- 3.6, compared to the mean MMSE score of 28.9 +/- 1.4 (Table 1). Conversely, patients with lower MMSE may be at a more advanced stage and thus have relatively more pronounced brain atrophy. Another possibility is that the specific cut-off values of MMSE and MoCA for cognitive impairment may have skewed the results. Furthermore, one of the limitations of our approach is that we used only the global measures of cognitive impairment; more detailed studies investigating correlations with domain-specific impairment (e.g. memory, attention, language, etc.) may provide further insight into how different cognitive assessments correlate with PET and MRI findings.

The study has several additional limitations. First, this study is a cross-sectional study using retrospective data. A key future study would be a longitudinal examination to determine how PET and/or atrophy positivity is associated with the progression of AD-related symptoms. It would also be of interest to determine whether PET-negative but atrophy-positive patients eventually become PET positive and/or become more cognitively impaired, compared to triple-negative individuals. Future studies may also incorporate additional clinical and genetic data, such as the APOE status and handedness, potentially with the use of artificial intelligence [32].

In the current study, there was a general trend of increased female representation correlated with PET positivity (% female = 50.7 for FBP-/FTP-, 54.1 for FBP+/FTP-, and 60.5 for FBP+/FTP+); more detailed examinations into the sex differences in the underlying brain structural and connectivity changes may give valuable insight into the pathogenesis and evolution of AD. More studies are also needed to determine whether elevated Aß and/or tau proteins directly cause brain regional atrophy and cortical thickness loss, as this study only examined correlations with a limited ability to draw causal conclusions. Participants were scanned on different scanners, which could introduce variability in the imaging acquisition. Third, the FBP-/FTP+ group had a very small sample size (n = 5), which limited our ability to analyze this group in depth. For instance, the small sample size precluded our analysis for associations with MMSE or MoCA since no one in the group fit the criteria for cognitive impairment using our cognitive test score cutoffs. Some of the MRI data, such as the white matter changes and vessels, were not included in the study. Lastly, this study did not examine any additional imaging-related and personnel costs associated with the triple-modality approach. It is likely that these additional costs will be variable depending on multiple factors such as location, insurance, etc., and will require a more comprehensive, multi-center study.

Taken together, our results demonstrate that cortical thickness loss and specific regional atrophy of the brain are associated with FBP or FTP positivity. The most prominent regional atrophy was observed in the temporal lobe, particularly in the entorhinal cortical thickness and overall temporal lobe GM volume. Double positivity in PET (FBP+/FTP+) was associated with greater regional atrophy and cognitive impairment. Triple positivity in both PET studies and MRI, particularly the temporal GM and entorhinal cortical atrophy, showed stronger associations with cognitive impairment than the PET studies alone. These findings show that a multimodality approach incorporating PET and MRI data may help improve the assessment of cognitive impairment in AD.

## Author contributions

All authors made substantial contributions to the study. Marlon Gonzales and Xiaojian Kang contributed to data collection, data analysis, and original draft preparation. Maheen M. Adamson and Steven Z. Chao contributed to study conceptualization as well as manuscript review and editing. Byung C. Yoon contributed to the study conceptualization, material preparation, data collection, funding acquisition, manuscript review and editing, and supervision.

## Data Availability

The data supporting the findings of this study are openly available via Alzheimer Disease Neuroimaging Initiative (https://adni.loni.usc.edu/).

## Acknowledgements

This study uses data from Data collection and sharing for this project was funded by the Alzheimer’s Disease Neuroimaging Initiative (ADNI) (National Institutes of Health Grant U01 AG024904) and DOD ADNI (Department of Defense award number W81XWH-12-2-0012). ADNI is funded by the National Institute on Aging, the National Institute of Biomedical Imaging and Bioengineering, and through generous contributions from the following: AbbVie, Alzheimer’s Association; Alzheimer’s Drug Discovery Foundation; Araclon Biotech; BioClinica, Inc.; Biogen; Bristol-Myers Squibb Company; CereSpir, Inc.; Cogstate; Eisai Inc.; Elan Pharmaceuticals, Inc.; Eli Lilly and Company; EuroImmun; F. Hoffmann-La Roche Ltd and its affiliated company Genentech, Inc.; Fujirebio; GE Healthcare; IXICO Ltd.; Janssen Alzheimer Immunotherapy Research & Development, LLC.; Johnson & Johnson Pharmaceutical Research & Development LLC.; Lumosity; Lundbeck; Merck & Co., Inc.; Meso Scale Diagnostics, LLC.; NeuroRx Research; Neurotrack Technologies; Novartis Pharmaceuticals Corporation; Pfizer Inc.; Piramal Imaging; Servier; Takeda Pharmaceutical Company; and Transition Therapeutics. The Canadian Institutes of Health Research is providing funds to support ADNI clinical sites in Canada. Private sector contributions are facilitated by the Foundation for the National Institutes of Health (www.fnih.org). The grantee organization is the Northern California Institute for Research and Education, and the study is coordinated by the Alzheimer’s Therapeutic Research Institute at the University of Southern California. ADNI data are disseminated by the Laboratory for Neuro Imaging at the University of Southern California. We would like to thank the Department of Statistics consulting service at Stanford University for guidance on statistical analysis.

## Ethical Considerations

This study only included data from a publicly available, de-identified database, Alzheimer’s Disease Neuroimaging Initiative (ADNI), which is approved by the Institutional Review Boards from the participating institutions.

## Consent to Participate

Informed consent specific to this study was not necessary because the current study only included data from a publicly available, de-identified database, Alzheimer’s Disease Neuroimaging Initiative (ADNI).

## Consent for Publication

The authors have given consent for the manuscript to be submitted in its current form.

