## Supplemental Tables for "Multimodal neuroimaging approach for cognitive impairment in Alzheimer’s disease"

**Supplemental Table 1. Anatomical lobes and Desikan-Killiany atlas parcels.**

| Lobes | Parcels |
| --- | --- |
| Frontal Lobe | Caudal middle frontal gyrus |
|  | Lateral orbital frontal gyrus |
|  | Medial orbital frontal gyrus |
|  | Paracentral lobule |
|  | Pars opercularis |
|  | Pars orbitalis |
|  | Pars triangularis |
|  | Precentral gyrus |
|  | Rostral middle frontal gyrus |
|  | Superior frontal gyrus |
|  | Frontal pole |
| Parietal Lobe | Inferior parietal gyrus |
|  | Postcentral gyrus |
|  | Precuneus cortex |
|  | Superior parietal gyrus |
|  | Supramarginal gyrus |
| Occipital Lobe | Cuneus cortex |
|  | Lateral occipital gyrus |
|  | Lingual gyrus |
|  | Pericalcarine cortex |
| Temporal Lobe | Banks superior temporal sulcus |
|  | Entorhinal cortex |
|  | Fusiform gyrus |
|  | Inferior temporal gyrus |
|  | Middle temporal gyrus |
|  | Parahippocampal gyrus |
|  | Superior temporal gyrus |
|  | Temporal pole |
|  | Transverse temporal gyrus |
| Cingulate Lobe | Caudal anterior cingulate gyrus |
|  | Isthmus cingulate gyrus |
|  | Posterior cingulate gyrus |
|  | Rostral anterior cingulate gyrus |
| Insula Cortex | Insular cortex |

**Supplemental Table 2. Variance inflation factors of the logistic regression analysis**

| **Cognitive impairment  (MoCA < 26 or MMSE <= 26)** | **Atrophied Region** | **VIF Amyloid** | **VIF Tau** | **VIF Atrophy** |
| --- | --- | --- | --- | --- |
| MoCA | Mean Cortical Thickness | 1.08 | 1.12 | 1.1 |
| MoCA | Entorhinal Cortex Thickness | 1.08 | 1.16 | 1.14 |
| MoCA | Temporal GM Volume | 1.09 | 1.12 | 1.58 |
| MoCA | Parietal GM Volume | 1.08 | 1.1 | 1.4 |
| MMSE | Mean Cortical Thickness | 1.11 | 1.17 | 1.13 |
| MMSE | Entorhinal Cortex Thickness | 1.1 | 1.24 | 1.26 |
| MMSE | Temporal GM Volume | 1.09 | 1.15 | 1.95 |
| MMSE | Parietal GM Volume | 1.11 | 1.13 | 1.7 |

*Variance inflation factors (VIFs) of amyloid-B, tau, and atrophy demonstrate relatively low (VIF < 2) correlations between the predictors.*

**Supplemental Table 3. Cortical thickness and GM volume changes by the FBP/FTP PET status.**

| **Region** | **FBP- FTP-** | **FBP+ FTP-** | **FBP- FTP+** | **FBP+ FTP+** | **R2** | ***p-*value** |
| --- | --- | --- | --- | --- | --- | --- |
| Mean Cortical Thickness | 2.372 | 2.364 | 2.300 | 2.282 | 0.073 | 3.72E-07 |
| **Cortical Thickness – LH** | | | | | | |
| Inferior Parietal | 2.318 | 2.308 | 2.206 | 2.161 | 0.119 | 1.24E-09 |
| Entorhinal | 2.887 | 2.741 | 2.538 | 2.511 | 0.108 | 1.65E-08 |
| Supramarginal | 2.401 | 2.402 | 2.230 | 2.279 | 0.102 | 2.5E-08 |
| Parietal Lobe | 2.244 | 2.243 | 2.146 | 2.123 | 0.1 | 7.4E-08 |
| Temporal Lobe | 2.677 | 2.640 | 2.528 | 2.515 | 0.097 | 8.23E-08 |
| Superior Temporal | 2.626 | 2.590 | 2.404 | 2.463 | 0.089 | 9.16E-08 |
| Precuneus | 2.295 | 2.285 | 2.250 | 2.154 | 0.09 | 1.61E-06 |
| Superior Parietal | 2.164 | 2.165 | 2.090 | 2.032 | 0.08 | 1.82E-05 |
| Middle Temporal | 2.675 | 2.658 | 2.596 | 2.532 | 0.066 | 0.000216 |
| Fusiform | 2.637 | 2.620 | 2.576 | 2.504 | 0.063 | 0.000395 |
| Lateral Occipital | 2.111 | 2.105 | 2.024 | 2.014 | 0.051 | 0.00337 |
| Parahippocampal | 2.611 | 2.577 | 2.532 | 2.408 | 0.047 | 0.00989 |
| Inferior Temporal | 2.646 | 2.634 | 2.484 | 2.537 | 0.046 | 0.017 |
| **Cortical Thickness – RH** | | | | | | |
| Temporal Lobe | 2.720 | 2.686 | 2.526 | 2.565 | 0.114 | 2.34E-09 |
| Entorhinal | 2.990 | 2.877 | 2.554 | 2.635 | 0.109 | 3.78E-08 |
| Superior Temporal | 2.651 | 2.616 | 2.442 | 2.514 | 0.083 | 6.89E-07 |
| Inferior Parietal | 2.359 | 2.351 | 2.308 | 2.228 | 0.082 | 6.23E-06 |
| Fusiform | 2.664 | 2.661 | 2.558 | 2.533 | 0.079 | 1.81E-05 |
| Precuneus | 2.320 | 2.296 | 2.290 | 2.202 | 0.075 | 0.000031 |
| Parietal Lobe | 2.251 | 2.251 | 2.198 | 2.148 | 0.071 | 4.66E-05 |
| Superior Parietal | 2.146 | 2.148 | 2.106 | 2.017 | 0.072 | 7.18E-05 |
| Middle Temporal | 2.711 | 2.695 | 2.640 | 2.587 | 0.061 | 0.00042 |
| Parahippocampal | 2.560 | 2.494 | 2.404 | 2.392 | 0.06 | 0.000561 |
| Supramarginal | 2.413 | 2.415 | 2.332 | 2.319 | 0.051 | 0.00322 |
| Banks Of The Superior Temporal Sulcus | 2.487 | 2.460 | 2.410 | 2.369 | 0.05 | 0.00521 |
| Isthmus Cingulate | 2.232 | 2.217 | 2.122 | 2.122 | 0.048 | 0.0134 |
| Lateral Occipital | 2.180 | 2.173 | 2.110 | 2.088 | 0.044 | 0.016 |
| Inferior Temporal | 2.675 | 2.673 | 2.546 | 2.586 | 0.04 | 0.0486 |
| **GM Volume – LH** | | | | | | |
| Temporal Lobe | 50297 | 48818 | 46526 | 44370 | 0.092 | 2.87E-14 |
| Inferior Parietal | 10829 | 10316 | 9685 | 9031 | 0.101 | 7.25E-10 |
| Middle Temporal | 9985 | 9597 | 9594 | 8546 | 0.08 | 1.34E-08 |
| Parietal Lobe | 50942 | 50422 | 49505 | 46305 | 0.055 | 9.39E-08 |
| Inferior Temporal | 10210 | 9813 | 9360 | 8835 | 0.067 | 3.45E-07 |
| Fusiform | 9072 | 8958 | 8824 | 8043 | 0.056 | 3.6E-06 |
| Superior Temporal | 11716 | 11373 | 10459 | 10578 | 0.054 | 1.64E-05 |
| Lateral Occipital | 11268 | 10693 | 11185 | 10145 | 0.055 | 3.05E-05 |
| Entorhinal | 1656 | 1585 | 1337 | 1361 | 0.066 | 5.58E-05 |
| Parahippocampal | 2021 | 1969 | 1822 | 1741 | 0.068 | 0.000137 |
| Precuneus | 8657 | 8560 | 8531 | 7823 | 0.044 | 0.000184 |
| Superior Frontal | 20621 | 20112 | 20532 | 19107 | 0.038 | 0.000593 |
| Frontal Lobe | 78664 | 77034 | 78325 | 75122 | 0.022 | 0.00933 |
| Superior Parietal | 12109 | 12053 | 12113 | 11112 | 0.029 | 0.0341 |
| **GM Volume – RH** | | | | | | |
| Temporal Lobe | 49550 | 48156 | 46116 | 44238 | 0.09 | 3.64E-14 |
| Middle Temporal | 10950 | 10651 | 11186 | 9541 | 0.076 | 1.61E-08 |
| Parietal Lobe | 52183 | 51340 | 51092 | 47472 | 0.056 | 5.41E-08 |
| Inferior Parietal | 13131 | 12732 | 11902 | 11403 | 0.075 | 1.06E-07 |
| Entorhinal | 1627 | 1543 | 1303 | 1313 | 0.08 | 2.39E-06 |
| Superior Temporal | 11055 | 10586 | 9945 | 10165 | 0.058 | 2.86E-06 |
| Inferior Temporal | 9893 | 9599 | 9130 | 8744 | 0.057 | 7.03E-06 |
| Parahippocampal | 1884 | 1800 | 1695 | 1653 | 0.074 | 2.56E-05 |
| Precuneus | 9132 | 8923 | 9115 | 8307 | 0.045 | 0.000103 |
| Superior Parietal | 11940 | 11793 | 11612 | 10769 | 0.048 | 0.000152 |
| Fusiform | 8882 | 8836 | 8008 | 8041 | 0.045 | 0.000366 |
| Frontal Lobe | 78535 | 77057 | 77313 | 74932 | 0.021 | 0.00807 |
| Banks Of The Superior Temporal Sulcus | 1969 | 1930 | 1802 | 1756 | 0.04 | 0.0112 |
| Superior Frontal | 19697 | 19238 | 19653 | 18475 | 0.027 | 0.0115 |
| Lateral Occipital | 11529 | 11184 | 11761 | 10619 | 0.029 | 0.0148 |
| Pars Triangularis | 3850 | 3664 | 4229 | 3581 | 0.038 | 0.0261 |
| Medial Orbitofrontal | 5338 | 5228 | 5290 | 5040 | 0.024 | 0.0459 |

*Table only includes cortical thickness and gray matter (GM) volume of brain regions with statistically significant group differences based on Bonferroni-adjusted p-values (*p *< .05). Cortical thickness measurements are in mm, and GM volume measurements are in mm^3^. RH, right hemisphere; LH, left hemisphere; FBP, florbetapir; FTP, flortaucipir; PET, positron emission tomography.*

**Supplemental Table 4. Incorporation of mean cortical thickness and regional brain atrophy to FBP/FTP for association with cognitive impairment**

| **Group (vs FBP- FTP-)** | **Group Size** | **OR** | **Lower 95% CI** | **Upper 95% CI** | **Bonf corrected *p*** | **Bonf corrected *p < .05*** |
| --- | --- | --- | --- | --- | --- | --- |
| **MMSE ≤ 26 - Mean Cortical Thickness** | | | | | | |
| FBP- FTP- Atrophy+ | 33 | 4.04 | 1.08 | 14.17 | 1.000 | N |
| FBP+ FTP- Atrophy- | 107 | 6.08 | 2.53 | 16.34 | 0.005 | Y |
| FBP+ FTP- Atrophy+ | 24 | 10.49 | 3.15 | 36.02 | 0.005 | Y |
| FBP+ FTP+ Atrophy- | 20 | 8.58 | 2.40 | 30.30 | 0.030 | Y |
| FBP+ FTP+ Atrophy+ | 17 | 28.47 | 8.35 | 105.32 | 0.000 | Y |
| **MoCA < 26 - Mean Cortical Thickness** | | | | | | |
| FBP- FTP- Atrophy+ | 33 | 1.19 | 0.53 | 2.67 | 1.000 | N |
| FBP+ FTP- Atrophy- | 107 | 1.69 | 1.01 | 2.85 | 1.000 | N |
| FBP+ FTP- Atrophy+ | 24 | 2.81 | 1.00 | 9.21 | 1.000 | N |
| FBP+ FTP+ Atrophy- | 20 | 3.14 | 1.10 | 10.49 | 1.000 | N |
| FBP+ FTP+ Atrophy+ | 17 | 20.05 | 3.80 | 371.18 | 0.181 | N |
| **MMSE ≤ 26 - Entorhinal Cortex, Cortical Thickness** | | | | | | |
| FBP- FTP- Atrophy+ | 28 | 9.45 | 2.64 | 34.19 | 0.018 | Y |
| FBP+ FTP- Atrophy- | 97 | 8.18 | 3.27 | 23.54 | 0.001 | Y |
| FBP+ FTP- Atrophy+ | 34 | 11.34 | 3.41 | 39.90 | 0.003 | Y |
| FBP+ FTP+ Atrophy- | 14 | 11.86 | 2.79 | 49.72 | 0.025 | Y |
| FBP+ FTP+ Atrophy+ | 23 | 27.21 | 8.35 | 97.24 | 0.000 | Y |
| **MoCA < 26 - Entorhinal Cortex, Cortical Thickness** | | | | | | |
| FBP- FTP- Atrophy+ | 28 | 1.83 | 0.78 | 4.47 | 1.000 | N |
| FBP+ FTP- Atrophy- | 97 | 1.59 | 0.94 | 2.71 | 1.000 | N |
| FBP+ FTP- Atrophy+ | 34 | 4.06 | 1.67 | 11.03 | 0.129 | N |
| FBP+ FTP+ Atrophy- | 14 | 2.33 | 0.71 | 9.09 | 1.000 | N |
| FBP+ FTP+ Atrophy+ | 23 | 15.07 | 4.03 | 99.11 | 0.020 | Y |
| **MMSE ≤ 26 - Temporal GM Volume** | | | | | | |
| FBP- FTP- Atrophy+ | 39 | 7.90 | 1.88 | 32.31 | 0.154 | N |
| FBP+ FTP- Atrophy- | 89 | 5.65 | 2.23 | 15.74 | 0.017 | Y |
| FBP+ FTP- Atrophy+ | 42 | 22.90 | 6.94 | 82.55 | 0.000 | Y |
| FBP+ FTP+ Atrophy- | 16 | 7.33 | 1.79 | 28.69 | 0.170 | N |
| FBP+ FTP+ Atrophy+ | 21 | 47.69 | 12.84 | 195.76 | 0.000 | Y |
| **MoCA < 26 - Temporal GM Volume** | | | | | |  |
| FBP- FTP- Atrophy+ | 39 | 2.05 | 0.88 | 4.85 | 1.000 | N |
| FBP+ FTP- Atrophy- | 89 | 1.86 | 1.07 | 3.27 | 1.000 | N |
| FBP+ FTP- Atrophy+ | 42 | 2.70 | 1.20 | 6.29 | 0.728 | N |
| FBP+ FTP+ Atrophy- | 16 | 5.23 | 1.51 | 24.81 | 0.684 | N |
| FBP+ FTP+ Atrophy+ | 21 | 8.29 | 2.46 | 38.37 | 0.074 | N |
| **MMSE ≤ 26 - Parietal GM Volume** | | | | | | |
| FBP- FTP- Atrophy+ | 37 | 6.99 | 1.77 | 26.57 | 0.167 | N |
| FBP+ FTP- Atrophy- | 100 | 6.28 | 2.56 | 17.09 | 0.005 | Y |
| FBP+ FTP- Atrophy+ | 31 | 17.79 | 5.03 | 66.93 | 0.000 | Y |
| FBP+ FTP+ Atrophy- | 20 | 10.62 | 3.07 | 37.29 | 0.007 | Y |
| FBP+ FTP+ Atrophy+ | 17 | 37.54 | 9.77 | 156.42 | 0.000 | Y |
| **MoCA < 26 - Parietal GM Volume** | | | | | | |
| FBP- FTP- Atrophy+ | 37 | 1.38 | 0.61 | 3.13 | 1.000 | N |
| FBP+ FTP- Atrophy- | 100 | 2.09 | 1.23 | 3.61 | 0.292 | N |
| FBP+ FTP- Atrophy+ | 31 | 1.32 | 0.53 | 3.33 | 1.000 | N |
| FBP+ FTP+ Atrophy- | 20 | 4.76 | 1.56 | 18.12 | 0.432 | N |
| FBP+ FTP+ Atrophy+ | 17 | 8.85 | 2.23 | 59.67 | 0.256 | N |

*The presence or absence of mean cortical thickness of both hemispheres or regional brain atrophy was added to the analysis of the associations between cognitive impairment (i.e. MMSE ≤ 26 or MoCA < 26) and the FBP/FTP status. FBP, florbetapir; FTP, flortaucipir; OR, odds ratio; CI, confidence interval; GM, gray matter; MMSE, Mini-Mental State Exam; MoCA, Montreal Cognitive Assessment; TIV, total intracranial volume; PET, positron emission tomography; Bonf corr, Bonferroni-corrected.*
